# Immunological presentations and clinical features associated with Thymic Malignancies: The potential role of histological classifications and tumour grading on the future recurrence of opportunistic infections and paraneoplastic autoimmune conditions

**DOI:** 10.1101/2024.07.29.24311199

**Authors:** Matthew A Abikenari, Sofia Delgado, Mohammad Ashraghi, Giacomo Greco, Andrew Tucker, Rhona Taberham, Mary Quirke, Dionisios Stavroulias, Slaveya Yancheva, Mark Mccole, Camilla Buckley, Maria Isabel Leite

## Abstract

Thymic malignancies are rare cancer tumours of the thymus arising from thymic epithelial cells and are characterized by a highly diversified clinical phenotype, substantial histologic and morphologic heterogeneity, and frequent presentations of associated paraneoplastic autoimmune syndromes. Myasthenia Gravis (MG) is the most prevalent of such autoimmune conditions, presenting in roughly half of thymoma patients, and is associated with substantial hyperactivation of T lymphocytes, highly dysregulated negative and positive T lymphocyte selection, leading to a systemic imbalance of the immune system, and consequently aiding and abetting the manifestation of severe opportunistic infections and multiple autoimmune comorbidities such as Pure Red Cell Aplasia and Good’s syndrome. Although the clinical, immunological and cytoarchitectural changes associated with thymomas have been increasingly elucidated in the contemporary literature, very few studies have interrogated the direct role of tumour staging and histological gradings on the occurrence and recurrence of infections and multiple autoimmune comorbidities. The current study aimed to interrogate the role of WHO thymoma classification criteria and Masaoka staging on the recurrence of severe opportunistic infections and the presentation of multiple paraneoplastic autoimmune syndromes post-thymectomy. The current study collected clinical and immunological data from 109 patients suffering from both MG and a pathologically proven thymoma. Statistical analysis of the collected data yielded significant associations between different stages of Masaoka grading and WHO classification on the number of autoimmune comorbidity and presence of severe recurrent infections, leading to the conclusion that early histological gradings and tumour infiltration patterns play a significant role in predicting future immunological behaviour, clinical outcomes, and susceptibility to recurrent infections. Future studies must further investigate the role of autoimmunity, its associated antibody expression profiles and thymic tissue pathology. Furthermore, novel therapeutics must further explore the role of emergent immunotherapeutics, such as adoptive cell therapies, as a viable patient-stratified treatment strategy for thymic malignancies.

## Introduction

The thymus is a lymphoid organ vital for the proper development, differentiation and proliferation of T lymphocytes which coordinate key adaptive immune responses throughout life. In addition, the thymus serves an essential role in instantiating immune self-tolerance and functions to prevent autoimmunity and substantial self-harm. Thymic malignancies are rare cancers of the thymus gland arising from tumours of thymic epithelial cells (TECs) and are associated with diverse clinical presentations, significant histologic and morphologic heterogeneity and unparalleled prevalence of associated paraneoplastic autoimmune diseases (Tomaszek et al., 2009; Shelly et al., 2011; Weksler and Lu, 2014). Thymic malignancies are extraordinarily fascinating and equally elusive to characterize due to the sheer spectrum of clinical and immunological presentations and respective multisystem disease features associated with the condition. Thymoma-the most frequent thymic tumour of the mediastinum-triggers substantial autoimmunity by releasing immature T lymphocytes into the systemic circulation and subsequently affecting multiple systems, organs and tissues in the body, leading to frequent opportunistic infections and additional paraneoplastic autoimmune syndromes (Tomaszek et al., 2009; Weksler and Lu, 2014). Myasthenia gravis (MG) is the most common of such autoimmune manifestations, a long-term neuromuscular junction disease, presenting in roughly 50% of patients with Thymoma (Ströbel et al., 2010; Romi, 2011a). However, only 15% of MG patients develop thymoma (Meriggioli and Sanders, 2012). Other associated autoimmune syndromes are rare in occurrence, present in roughly 1-5% of patients per illness and include pure red cell aplasia (PRCA), parathyroid adenoma, hypogammaglobulinemia (Good Syndrome), autoimmune hepatitis, syndrome of inappropriate antidiuretic hormone secretion (SIADH), and various other paraneoplastic syndromes (MORGENTHALER et al., 1993; Engels, 2010). Thymomas follow an unpredictable natural history and disease progression, ranging from clinical cases characterized by the presence of aggressively malignant tumours to cases with an indolent disease course, including long asymptomatic phases and the accidental discovery of the disease from a chest X-ray/CT scan related to another condition (Pelosof and Gerber, 2010; Romi, 2011a). Although the exact aetiology and oncogenesis of thymomas remain inconclusive, there has been substantial progress over the last few decades in understanding the cellular, molecular and aberrant pathways involved in the pathogenesis of thymoma and its associated paraneoplastic autoimmunity (Ströbel et al., 2010; Soomro, 2020).

### Epidemiology

Thymoma and associated malignancies are extremely rare in occurrence, with an overall incidence rate of 0.13-0.31 per 100,000 people per year, accounting for roughly 03.-1.5 of all malignancies (de Jong et al., 2008; Engels, 2010). The European crude incidence rate of malignant thymoma was 1.4 per million yearly. There are, on average, 1000 cases of thymoma per year in Europe, measured by the RARECAREnet projects that focus on the prevalence of rare cancers in Europe (Siesling et al., 2012a; Kaneko et al., 2018; Rich, 2020). The majority of the literature indicates that sex has no influence on the development of the disease, and the incidence of thymic malignancies presents equally for men and women (de Jong et al., 2008; Margaritora et al., 2010; Siesling et al., 2012a, 2012b). The mean age group associated with the peak incidence of thymic malignancies appears to be in the middle age of 45-55 (Engels and Pfeiffer, 2003), although a large population study by the EUROCARE-4 network of cancer registries in Europe reported a mean age of presentation of 56 years (Siesling et al., 2012a; Rich, 2020). Thymoma is extremely rare in children and young adults, although cases have been reported in every age group (Engels, 2010; Rich, 2020). One fascinating observation about the incidence of thymoma is that although the increase in rates of malignancy with increasing age is in line with the age-related build-up of the disease (Engels, 2010; Rich, 2020), it is surprising that this is the case in thymic malignancies as the thymus gland itself gets substantially reduced in size with increasing age.

### Clinical Presentation, Staging and Treatment

Clinical manifestation of thymoma can vary depending on disease severity, associated conditions and pace of progression. However, the majority of patients present clinically in one of the three following ways: First, a third of thymoma patients will have no symptoms, and their diagnosis is accidentally discovered by a CT/CAT scan or a chest X-ray for an unrelated medical reason. A third of patients are diagnosed with thymoma due to the presence of associated paraneoplastic autoimmune syndromes such as MG, Good Syndrome, PRCA or various other autoimmune comorbidities. The final third of thymoma patients are diagnosed due to severe symptoms arising from the compression of surrounding organs of the chest by the presence of the thymic tumour (Masaoka et al., 1981; Kim and Thomas, 2015; Bernard et al., 2016a). The enlargement of the thymic tumour can lead to encroachment and obstruction of the superior vena cava, leading to superior vena cava syndrome (SVCS) and resulting symptoms such as shortness of breath, chest pain, difficulty swallowing, oedema and chronic coughing (MORGENTHALER et al., 1993; Romi, 2011b).

Thymoma staging and classification are commonly done at the time of surgery, using the Masaoka Staging System (Figures 1 and 2), based on the degree of tumour infiltration at the time of operation. Stage I is defined as the lack of capsular invasion. Stage IIA is defined as a microscopic invasion of the capsule. Stage IIB is defined as the macroscopic invasion of the surrounding fatty tissue or the mediastinal pleura. Stage III is a macroscopic invasion of the surrounding organs. Stage IVA is defined as pericardial dissemination, and Stage IVB is defined as hematogenous or lymphatic dissemination to distant sites (30). On average, five-year overall survival outcomes in each of the clinical stages of treated thymoma patients appear as follows: 93% in stage I, 85% in stage II, 69% in stage II, and 50% in stage IV. 10-year overall survival rate ranges from 54-65% in treated patients (Masaoka et al., 1981; Verley and Hollmann, 1985).

**Figure 1:**
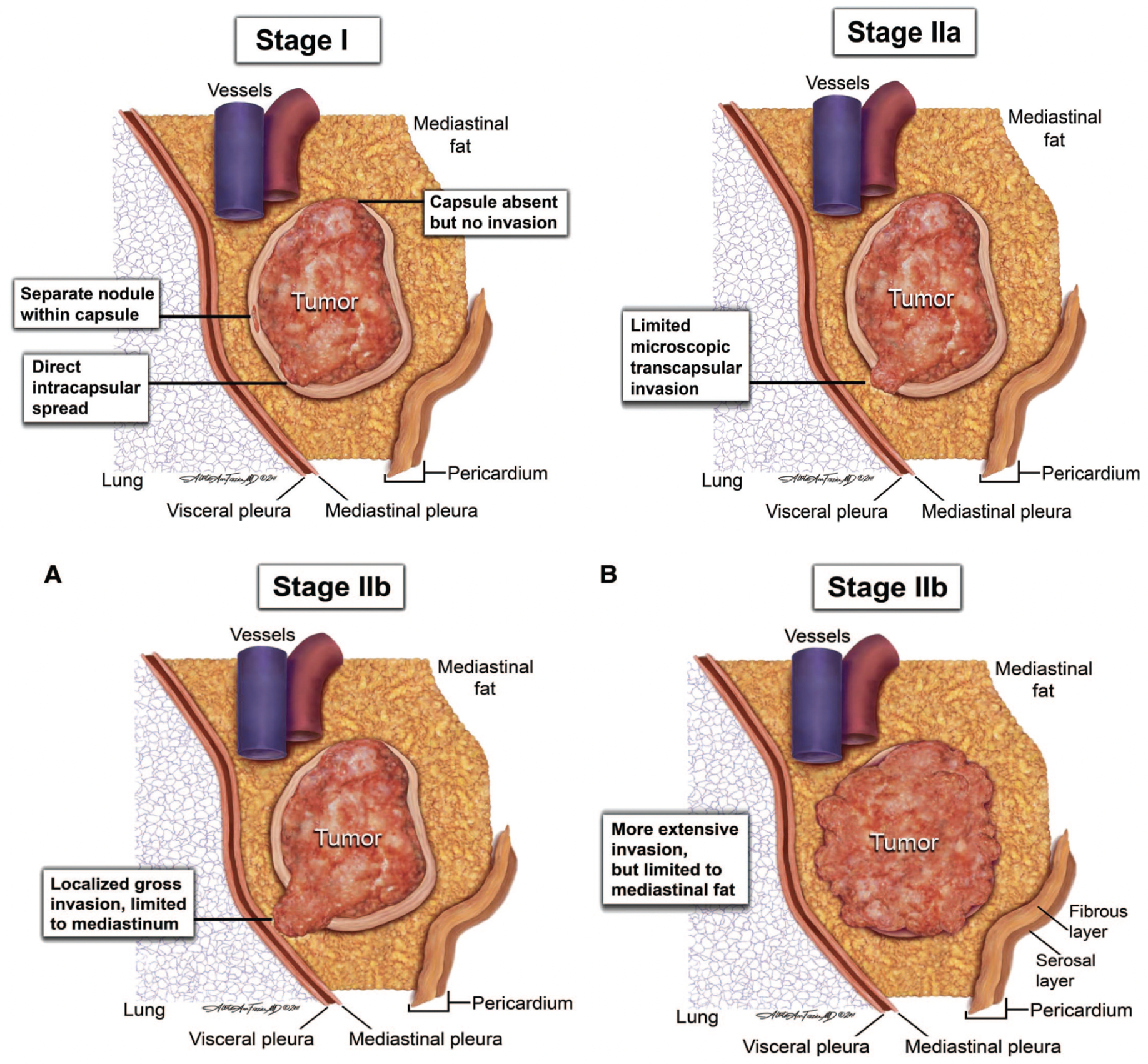
Masaoka-Koga Staging System and Schematics of tumour infiltrations associated with thymic malignancies. Stage I: Penetrations within the capsule are considered noninvasive in thymoma pathology, although a partial invasion of the capsule exists. Stage IIa: Schematic of microscopic transcapsular invasion. Stage IIb: Both A and B types of stage IIb are characterized by capsule invasion; however, they range from single areas of localized invasion (A) to extensive invasion of the mediastinal fat without invasion into the neighbouring organs. Figure 1 adapted from (Detterbeck et al., 2011)

**Figure 2:**
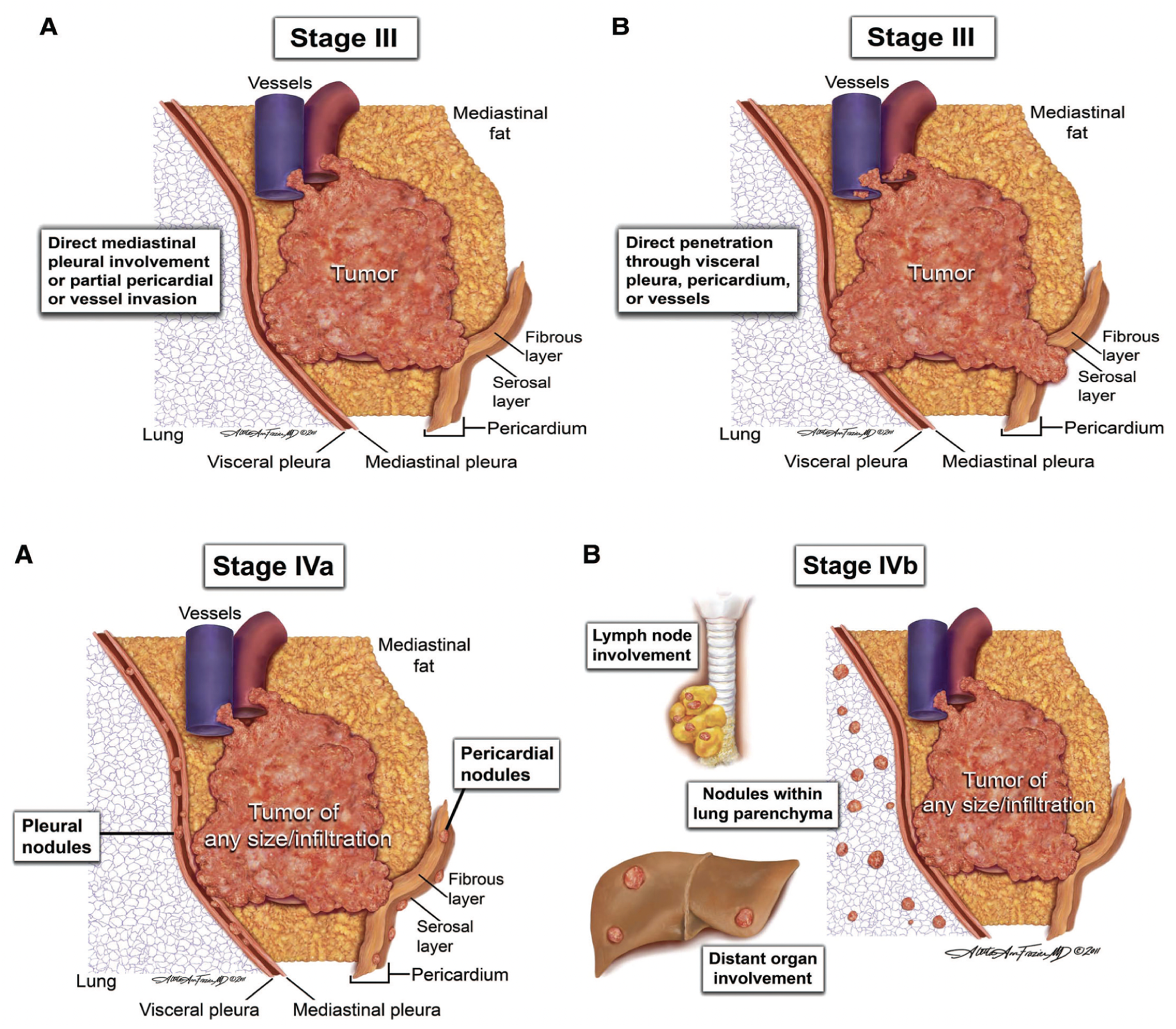
Masaoka-Koga Staging System and schematics of tumour infiltrations associated with thymic malignancies continued. Stage III: Both A and B types of stage III are characterized by capsule invasion and macroscopic invasion of the neighbouring organs, such as involvement of mediastinal pleura, pericardium, the visceral pleura, the phrenic nerve, and invasion through major vascular structures. Stage IIIB is characterized by the invasion through the pericardium. Stage IV: Both A and B types of stage IV are characterized by tumour infiltration across the pericardium. Stage IVA involves pleural or pericardial metastases. Stage IVB involves lymphogenous and hematogenous metastasis and distal metastases outside the cervical perithymic region. Figure 2 adapted from (Detterbeck et al., 2011)

**Figure 3:**
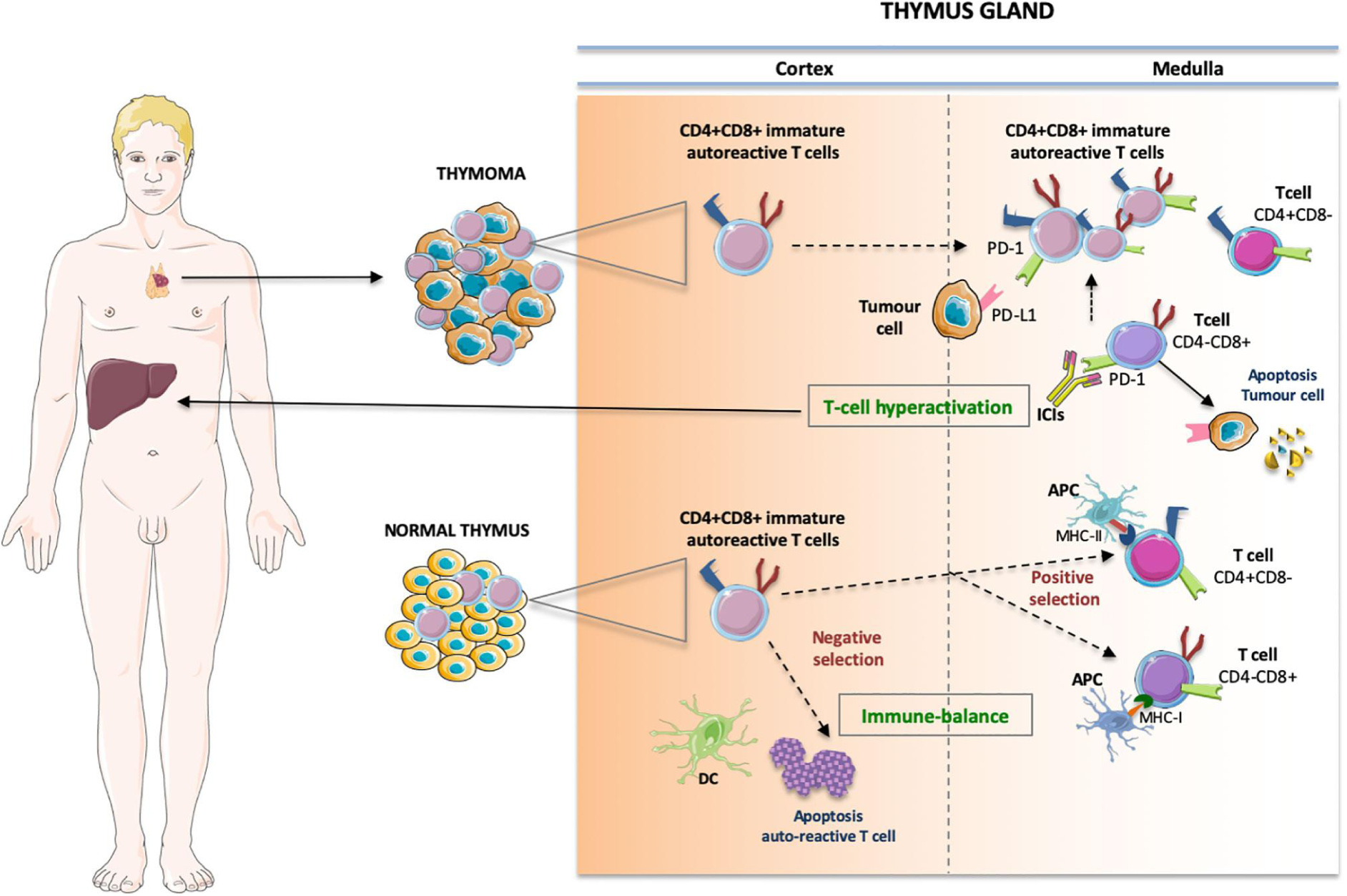
The Immunological cascade is frequently observed in the pathophysiology of thymic malignancies and associated paraneoplastic presentations. The vicious immunological cascade includes enhanced T-Lymphocyte activation and proliferation against antigens present in the thymus and surrounding organs. The pre-existence of large populations of autoreactive T cells released from the thymic medulla breaks the thymic immune homeostasis and further induces dysregulation of thymic immunity. Specifically, the negative selection causes apoptosis of self-reactive T clones, and immature double-positive CD4+ and CD8+ cells migrate to single positive CD8+ cytotoxic T cells and CD4+ helper T cells, binding to MHC class I and II, respectively. Through dendritic cell-prompted T-lymphocyte hyperactivation and autoimmune presentations, immature thymic lymphocytes escape quality control and further perpetuate the vicious cycle of autoimmunity. Figure 3 adapted from (Argentiero et al., 2020)

Thymectomy-complete surgical resection of the thymus gland-is the principal approach for treating thymoma (Rea et al., 2004). Despite successful resection of the thymic tumour, roughly 15-30% of patients experience recurrent thymoma, with a disease-free phase of approximately 60-80 months (Rea et al., 2004; Marulli et al., 2017). In patients with late-stage Masaoka/WHO histology classification, neoadjuvant radiotherapy and chemotherapy are frequently used to reduce the size and enhance the resectability odds of the tumour prior to thymectomy. The primary chemotherapeutic strategy for thymoma patients includes the combined administration of cisplatin, doxorubicin, and cyclophosphamide (referred to as CAP) (Thomas et al., 1999; Berghmans et al., 2018). Unsurprisingly, thymoma patients suffering from the associated autoimmune syndrome of MG are given immunosuppressants such as azathioprine (AZA), rituximab (RTX), or methotrexate (MTX) to lower the highly pronounced autoimmune activity (Kumar and Kaminski, 2011; Gotterer and Li, 2016; Tandan et al., 2017).

Contemporary oncological advances in the treatment of thymic malignancies have primarily focused on the role of targeted therapies such as tyrosine kinase inhibitors (TKIs) and mammalian target of rapamycin (mTOR) inhibitors as well as immunotherapies such as antibody-engineered immune checkpoint inhibitors (ICIs) and tumour-associated antigens (TAAs) cancer vaccines (Martin et al., 2015; Zhang and Chen, 2018; Ballman et al., 2022a). It is important to note that although there has been a greater interrogation by contemporary studies on the role of autoimmunity, antibody-induced ICIs and multisystem disease features of thymoma (Velcheti and Schalper, 2016; Lippner et al., 2019; Ballman et al., 2022b), very few studies have examined the precise influence of autoimmune presentations, tumour histology and antibody profiles on the occurrence of opportunistic infections and patient-stratified clinical phenotype.

### Pathophysiology, autoimmunity, and systemic vulnerability

Thymoma tumours arising from TECs are highly heterogeneous in morphology and histology. Type A thymomas contain neoplastic cells with oval-shaped or spindle nuclei and few non-neoplastic lymphocytes. Type B thymomas are characterized by a significant presence of immature lymphocytes, and they mimic the appearance of a normal thymic cortex. Type AB thymomas resemble the combined characteristics of both type A and B thymomas (Weksler and Lu, 2014; Marx et al., 2015). Unlike the Masaoka system, which is based on the behaviour of the tumour and capsule infiltration of neighbouring structures, the WHO classification (Table 1) focuses on microscopic histological criteria in order to differentiate thymic tumour type (Kondo et al., 2004; Kondo, 2008; Shelly et al., 2011).

**Table 1.** WHO classification of Thymoma.

|  |  |
| --- | --- |
| **A** | Thymic tumor with an oval shape, absence of both nuclear atypia and non-neoplastic lymphocytes |
| **AB** | Thymic tumor with foci resembling characteristics of thymoma type A mixed with the presence of lymphocyte rich foci |
| **B1** | Thymic tumor resembling characteristics of a normal and functional thymus, with an expansive appearance of normal thymic cortex and thymic medulla |
| **B2** | Thymic tumor in which neoplastic epithelial cells are present in the shape of scattered cells with vesicular nuclei and distinct nucleoli saturated in a large pool of lymphocytes |
| **B3** | Thymic tumor characterized by the predominance of epithelial cells with round and polygonal shapes and displaying no nuclear atypia |
| **C** | Thymic tumor characterized by the absence of immature lymphocytes, a clear sign of cytologic atypia, and the presence of features of no longer unique to thymic malignancies but rather features comparable to carcinomas of other organs |

Given that the thymus is essential for T lymphocyte maturation and development, it is unsurprising that thymomas are heavily linked with many paraneoplastic autoimmune manifestations. An intriguing feature of thymoma pathophysiology is that they generate T lymphocytes that mature into CD4+ and CD8+ lymphocytes (Buckley et al., 2001; Weksler and Lu, 2014). Furthermore, each WHO thymoma type carries a differentiated pattern of thymopoiesis and consequent negative and positive T lymphocyte selection. Type B thymomas generate substantially greater lymphocytes than type A and are most associated with autoimmune conditions. This is due to the unique cytoarchitectural features of type B thymomas which exhibit a heavily disorganized cortex and contain neoplastic epithelial cells that display virtually no major histocompatibility complex class II molecules (MHC II), which is essential for T lymphocyte positive selection (Weksler and Lu, 2014; Tateo et al., 2020; Ballman et al., 2022a).Consequently, such impaired regulation of positive selection leads to systemic deprivation of functional and responsive T lymphocyte repertoire.

Furthermore, the generation of regulatory T lymphocytes (Tregs), which serve a critical role in the supervision of cytokine and lymphocyte proliferation, self-tolerance and prevention of autoimmunity, are compromised in thymoma patients (Weksler and Lu, 2014; Jamilloux et al., 2018). Impaired negative selection is another key characteristic in the pathophysiology of thymomas. The majority of neoplastic cells in thymoma fail to express the AIRE transcription factor, a critical autoimmune regulator protein in T lymphocyte negative selection (Ströbel et al., 2010; Weksler and Lu, 2014; Girard, 2019; Conforti et al., 2020). Negative selection is a vital step for establishing immune tolerance, as it eliminates potentially self-reactive thymocytes and enables the immune system to generate peripheral supplies of self-tolerant T lymphocytes and develop autoimmune tolerance (Cheng and Anderson, 2018; Zhao and Rajan, 2019). It is important to note that impaired regulation of negative self-selection in thymoma patients is particularly consequential as they permit the release of autoreactive T lymphocytes, which are key pathologic players in autoimmune conditions (Bernard et al., 2016b; Zhao and Rajan, 2019; Ballman et al., 2022a). It is plausible that this particular cascade may predispose patients to the numerous paraneoplastic autoimmune manifestations characteristic of thymoma.

The most frequent of such autoimmune manifestations, MG, is particularly insightful in guiding our understanding of dysregulated autoimmunity. It is least surprising that nearly half of thymoma patients present with MG, given the overwhelming overlap in their underlying immunological behaviour and antibody expression profiles (Mao et al., 2012; Weksler and Lu, 2014; Ballman et al., 2022a). For instance, MG patients without thymoma share numerous antibodies with MG patients with thymoma (Marx et al., 2010; Decroos et al., 2014). Both types of patients frequently present with anti-striatal antibodies (StrAbs), immunoglobulins known to react against muscle antigens, particularly ryanodine and titin receptors (Mckeon et al., 2013; Decroos et al., 2014). Furthermore, both MG and thymoma patients share parallel cytokine, interferon, and acetylcholine receptor (AChR) autoantibodies. The antibody expression profiles of the two patient types are so similar that the presence of thymoma-specific antibodies in early-onset MG patients can serve as a prediction model for the presence of thymic malignancy (Marx et al., 2003; Mao et al., 2012; Decroos et al., 2014; Weksler and Lu, 2014).

Although there has been substantial progress over the last two decades in the elucidation of underlying mechanisms driving pathology in thymoma and associated paraneoplastic syndromes (Burbelo et al., 2010; Evoli and Lancaster, 2014; Tateo et al., 2020; Ballman et al., 2022a), many of such mechanisms fail to provide a consistent prediction model of clinical phenotype based on the associated immunological presentations. For instance, thymoma patients with type B1 tumours follow the appropriate expression of AIRE protein, yet they experience an overwhelming degree of associated autoimmune conditions (Girard, 2019).

Moreover, patients with thymoma exhibit clinical phenotypes and antibody expression profiles that differ from patients who display similar paraneoplastic autoimmune conditions purely as a result of faulty AIRE expression (Lippner et al., 2019; Ballman et al., 2022a). Furthermore, despite the overwhelming overlaps in clinical phenotype, autoimmune presentations and antibody expression profiles of patients with MG and thymoma, few clinical studies to date have interrogated such underlying parallels in autoimmunity, recurrent infections and clinical profiles frequently present in patients suffering from both MG and thymoma. Lastly, no studies to date have investigated a potential prediction model of recurrent infections and autoimmune presentation based on WHO and Masaoka grading classifications.

### Objectives of the current study

Given that patients with thymoma may develop one or more autoimmune conditions associated with thymic recurrence, opportunistic infections and a highly heterogeneous clinical phenotype, establishing the frequency and type of autoimmune features, associated antibody profiles and recurrent infections with thymic pathology and disease severity will provide us with a greater understanding of the relationship between thymic malignancy and autoimmunity, and subsequently aid in the potential development of prediction models of clinical outcomes and patient-stratified treatment strategies that lead to the early diagnosis and treatment of thymomas. The current study aimed to (i) establish the frequency and spectrum of immunological presentations and clinical phenotype present in patients suffering from both thymic malignancy and myasthenia gravis; (ii) investigate the occurrence and recurrence of opportunistic infections and autoimmune conditions with disease stage and histological classifications; (iii) explore whether there is a link between mortality and the type of thymic tumour, clinical phenotype and multisystem manifestations (autoimmune or infections) in thymoma patients suffering from at least one autoimmune paraneoplastic condition.

## Methods

### Participants

Given the highly rare occurrence of thymic malignancies in Europe, the analyzed data in the current study was collected over five decades and included frequent follow-ups of some patients for more than 30 years, providing a rich clinical and immunological data set, uncommon in the current thymoma literature. The current study included patients with a pathologically proven thymoma who underwent thymectomy in Oxford from 1970 to January 1st 2023. All patients had a concomitant diagnosis of paraneoplastic autoimmune syndrome of Myasthenia Gravis (MG). All patients included in the current study underwent treatment at John Radcliffe Hospital and associated Oxford University Hospitals and were part of the Nuffield Department of Clinical Neurosciences Medical Sciences Division. All patient data were collected from hospital medical charts. All patients in the current study signed an informed consent for their data to be collected for research purposes (REC reference 16/YH/0013). All procedures and collected clinical data were approved by the Leeds East Research Ethics Committee and the University of Oxford’s Central University Research Ethics Committee. The analyzed data for the current study was derived from a sample of 109 patients (37 Male, 72 Female), on whom the appropriate clinical criteria for inclusion into the study were met (Table 2). The age of patients ranged between 31 and 102 years (mean = 65.90 ± 15.87 years). The mean age at which patients of the current study underwent thymectomy ranged between 16 and 98 years (mean = 49.34 ± 15.49 years). The number of years each patient was followed up throughout the study ranged between 1 and 33 years (mean = 10.10 ± 8.40 years). Given the decades-long length of the current study, and the fact that many patients who underwent thymectomy at Oxford were not first diagnosed in the United Kingdom, the distribution and mean age of diagnosis of patients in the current study were not included.

**Table 2.** Patient’s demographic information.

|  | *N(%)* | *Mean (SD)* | *Range (min - max)* |
| --- | --- | --- | --- |
| Patient sample of study | 109 |  |  |
| Gender |  |  |  |
| Female | 72 (66%) |  |  |
| Male | 37 (34%) |  |  |
| Age |  | 65.90 (15.87) | 31 - 102 |
| Age at thymectomy |  | 49.34 (15.49) | 16 - 98 |
| Follow-up years |  | 10.10 (8.40) | 1 - 33 |

*Note.* SD = standard deviation

## Data Analysis

Prior to statistical analysis, all of the clinical data used in the study was visually inspected through Excel to ensure there were no missing cells or partial patient data and no incorrect entries of clinical data derived from patient medical records. The entirety of the statistical analysis in the current study was performed using SPSS software version 28.0 (IBM SPSS Statistics, NY, USA). Existing data were checked for outliers, heteroscedasticity, and residual normality using SPSS. Robust regression models were used to detect and exclude any outliers in the data set. Outliers were defined as any variable in the data set greater or less than three times the interquartile range. Given the clinical target of the current study, all included thymoma patients had an accompanying diagnosis of myasthenia gravis (MG). Hence, thymoma patients without a clinical diagnosis of myasthenia gravis were excluded from the dataset, which included a total of 10 patients.

The statistical analysis included comparisons of demographic variables with clinical variables and inter and intra-group clinical comparisons. Demographic variables included age, sex and age of thymectomy. Clinical variables included WHO histological classification, Masaoka grading scale, presence of MG, paraneoplastic autoimmune comorbidities, presence and profiles of autoantibodies, immunosuppressive treatments, infections, recurrent infections, recurrent thymoma and hypogammaglobulinemia. Statistical differences between levels of independent clinical variables were analyzed using a parametric one-way factorial analysis of variance (ANOVA) to allow for the comparison of multiple means of clinical criteria at once. The F test was used for statistical significance to compare the variance in each group’s mean with the overall variance. Statistical differences between clinical criteria for the same subset of patients were analyzed using a parametric dependent samples t-test. This was particularly important for the current study as the project aimed to elucidate clinical and immunological manifestations across the same subset of patients. Furthermore, a dependent samples t-test ensures unrelated variation in clinical phenotype is eliminated and reduces sampling error for the differences in means of clinical categories of interest.

To identify the number and spectrum of clinical and immunological conditions associated with thymic malignancies, the current study performed descriptive statistics to derive the frequency of associated paraneoplastic autoimmune conditions such as pure red cell aplasia, good’s syndrome, thyroid disease, autoimmune hepatitis, syndrome of inappropriate secretion of antidiuretic hormone (SIADH) and autoimmune skin disorders. Furthermore, the overall frequency of infection, recurrent infection and hypogammaglobulinemia in the respective patients were also derived into the descriptive statistics to shed light on the prevalence of opportunistic infections during autoimmune presentations. In order to investigate the occurrence and recurrence of opportunistic infections in thymoma patients with single or multiple autoimmune comorbidities, a linear regression analysis was derived to depict the relationship between the presence of autoimmune comorbidity and the occurrence of opportunistic infections.

Furthermore, to explore the link between the type of thymic tumour, mortality and occurrence of infections and autoimmune presentations, a two-way factorial analysis of variance (ANOVA) was performed to determine if the level of WHO classifications and Masaoka staging affected the occurrence and recurrence of opportunistic infections and autoimmune presentations. A post hoc dependent-samples t-test followed ANOVA analysis to determine the presence of any simple effects in the analysis. Statistically significant results in the current study were defined as analyses with a *p*-value of 0.05 or lower.

## Results

The demographics, clinical variables and descriptive statistics of associated clinical phenotypes for the patient data set are described in **Table 3**. Although the current study has a predominantly female sample of thymoma patients (72 Females, 37 Males), there was no statistically significant difference in the prevalence of associated autoimmune comorbidity and recurrent infections between the male and females of the study (*p*-value = 0.505). Moreover, there was also no statistically significant difference between the genders and the type of thymic tumour (WHO classification and Masaoka staging) (*p*-value = 0.073) or between the genders and the presence of thymic hyperplasia in the remaining tissue (*p*-value = 0.061).

**Table 3.**
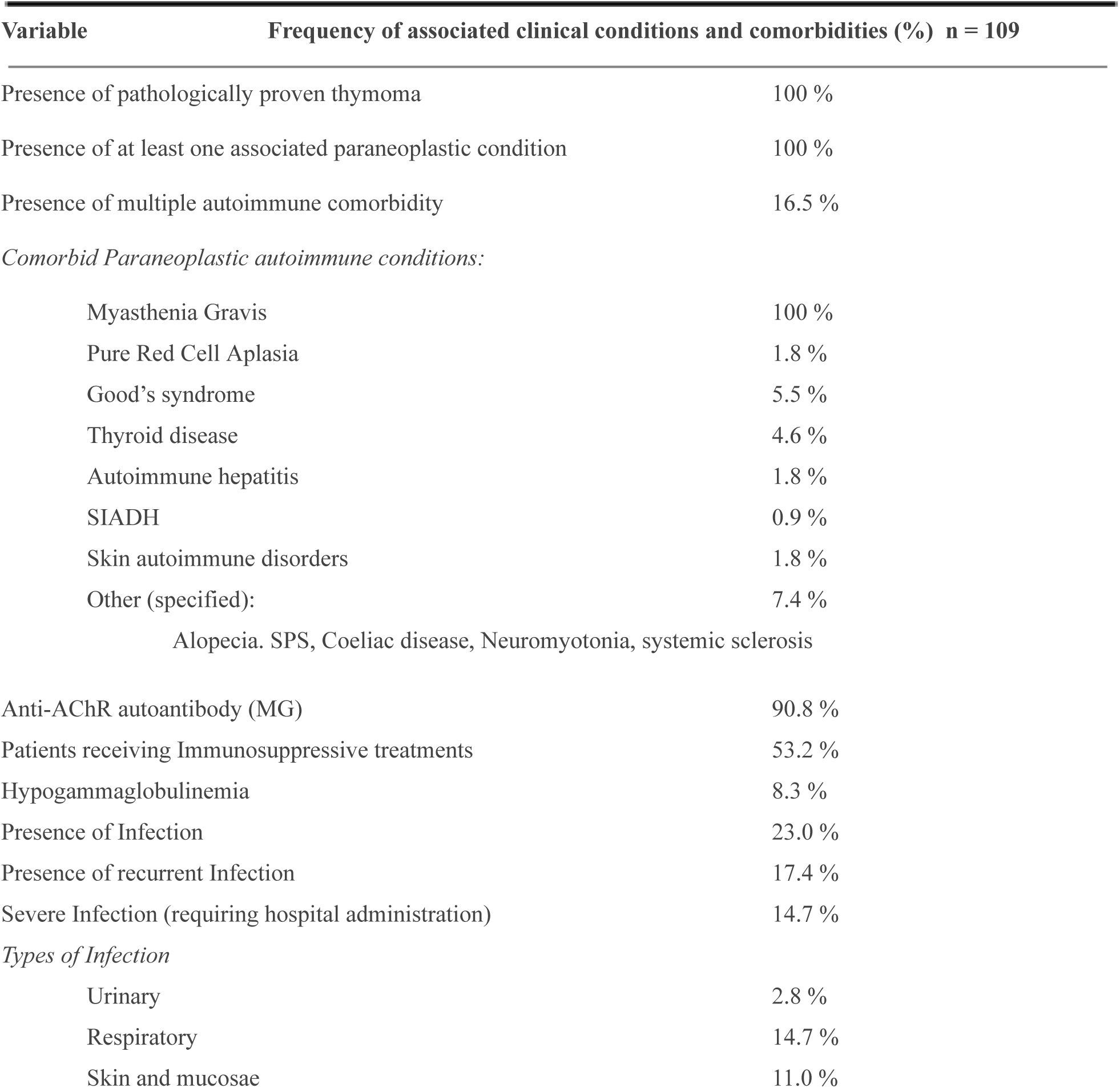
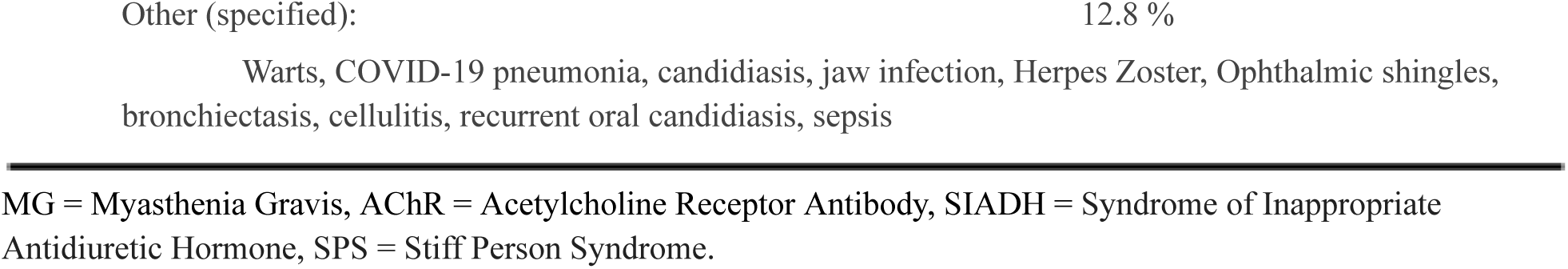
Patient demographics and frequencies of clinical and immunological phenotypes in the current study sample.

The one-way factorial analysis of variance (ANOVA) of the effect of different stages of Masaoka grading and WHO classification on the number of autoimmune comorbidities yielded no statistically significant difference in the means across various histological gradings, *F(*1, 99) = 2.11, *p* = .150 (WHO), *F(*1, 88) = 3.03, *p* = .085 (Masaoka). Therefore, high-grade tumour histology presented no predictive utility or statistical association with the number of comorbid autoimmune presentations.

An additional one-way factorial analysis of variance (ANOVA) on the effect of different stages of Masaoka grading and WHO classification on the age of thymoma-diagnosed patients also yielded no statistically significant difference across the various histological gradings, *F(*1, 100) = 1.17, *p* = .289 (WHO), *F(*1, 88) = .856, *p* = .694 (Masaoka). Therefore, high-grade tumour histology presented no predictive utility or statistical association with increased age in patients diagnosed with thymic malignancies. The dependent samples t-test on the effect of different histological classifications early in the diagnosis and the presence of recurrent opportunistic infections yielded a statistically significant difference, in particular on the paired association of WHO classification and recurrent infection (M= 2.34; SD= 1.21); [t(100) = 19.43, *p* < .001.

The paired association of Masaoka staging and recurrent infection yielded no statistically significant difference between the means (M= 1.31; SD= .876); [t(85) = .985, *p* = .328. Unlike the WHO classification, severity in Masaoka staging at the time of diagnosis did not affect the future recurrence of opportunistic infection.However, a dependent samples t-test of severity in Masaoka staging prior to thymectomy yielded a significant paired effect with the presentation of multiple autoimmune conditions after thymectomy (M= .955; SD= .891); [t(88) = 10.11, *p* < .001.Bivariate Pearson correlations of the clinical and immunological phenotypes in the current study sample revealed a moderate and statistically significant relationship between the occurrence of opportunistic infection and the presence of multiple paraneoplastic autoimmune conditions (*r*=.373, *p* < .001). Furthermore, there was a moderate and statistically significant association between recurrent infection and the presence of multiple paraneoplastic autoimmune conditions (*r*=.306, *p* = .001). Lastly, there was a moderately significant association between WHO classification and Masaoka staging (*r*=.313, *p* = .003).

## Discussion

The current study sought to determine first the frequency and spectrum of immunological presentations, opportunistic infections, clinical phenotype and multisystem disease features associated with thymic malignancies. Second, the current study aimed to investigate whether there is an association between the type of thymic tumour (defined by WHO histological classification and Masaoka staging) and the occurrence and recurrence of opportunistic infections and comorbid paraneoplastic autoimmune conditions in patients suffering from pathologically proven thymomas. Third, the current study aimed to explore whether predictive associations exist between histological data and autoimmune comorbidity and infection severity. The derived analysis of the current project resulted in a few key findings. First, the current study elucidated the frequency and subtype of paraneoplastic autoimmune conditions and opportunistic infections in a large sample of pathologically proven thymoma patients that were characterized by highly heterogeneous tumour histology and clinical phenotypes. Second, the current analysis revealed a statistically significant association between WHO classifications of tumour pathology and the recurrence of opportunistic infections post-thymectomy. Furthermore, the current study demonstrated a statistically significant association between increasing infiltration of thymic tumour based on the Masaoka staging and the increase in manifestations of paraneoplastic autoimmune syndromes after thymectomy. Bivariate Pearson correlations of all immunological and clinical criteria corroborated the major findings, as there was a statistically significant relationship between the occurrence and recurrence of sub-severe and severe opportunistic infections and the increase in autoimmune comorbidities. The driving emphasis of the current study was to elucidate the predictive nature of tumour histology using WHO classification criteria and microscopic and macroscopic signatures of tumour infiltration using Masaoka staging. The study also revealed a complementary relationship between the two grading systems, as there was a statistically significant association between the thymic tumour WHO classification and Masaoka stagings assigned to the patient sample in the current study.

Although the current study demonstrated the frequency and spectrum of clinical and immunological criteria associated with the disease, as well as clear links between thymic tumour type and occurrence of autoimmune comorbidity and opportunistic infections, the analysis failed to demonstrate predictive associations between patient demographics, autoimmune expression profiles, tissue pathology and subsequent disease severity. In particular, the derived analysis did not reveal any statistically significant association between increased age and an increase in tumour tissue grade and pathology. Furthermore, although the predominant lens through which associations were derived in the current study was thymic tumour type using WHO classifications and Masaoka staging, the current study failed to demonstrate synergistic associations by combing WHO classification and Masaoka staging to predict multisystem disease features associated with thymic malignancies.

Unlike many of the contemporary clinical studies, which focus exclusively on patients diagnosed with thymic malignancies or Myasthenia Gravis (MG), the current study aimed to elucidate the clinical and immunological presentations of patients suffering from both MG and pathologically proven thymomas. This is a particularly insightful selection criterion as the systematic presence of MG in all of the patients in the current study serves as a control variable in highlighting immunological behaviours not attributed solely to MG and aids in illuminating the influence of other highly involved paraneoplastic conditions such as Pure Red Cell Aplasia, Good’s Syndrome and SIADH. Furthermore, the data collected in the current study is particularly insightful on the nature of autoimmune comorbidities frequent in thymic malignancies as it interrogates the occurrence and recurrence of multisystem disease features in the same set of patients across decades, shedding light on the highly heterogeneous disease trajectory of thymoma patients.

The presence of multiple paraneoplastic autoimmune conditions in roughly 1 in 6 thymoma patients of the current study (Table 3) is consistent with the literature (Evoli and Lancaster, 2014; Solimani et al., 2019; Zhao et al., 2020), highlighting the profound influence of autoimmunity in driving the clinical phenotypes associated with the disease. Furthermore, despite overwhelming parallels in diagnosis, clinical phenotypes, and equivalent WHO and Masaoka histological gradings with similar demographics, patients in the current study displayed highly heterogeneous autoimmune presentations and opportunistic infections. Although the autoimmune features and infection severity of the patients were highly diversified, the trajectory and phases in immune vulnerability (post-thymectomy, hospitalizations etc.) were highly similar, alluding to the fact that emergent therapeutics in thymomas must address underlying mechanisms in susceptibility to immune imbalance instead of addressing emergent clinical presentations as a consequence of the underlying autoimmunity. For instance, it is well characterized in the literature that many patients with thymoma will clinically present with an immunodeficiency disorder while they are suffering from an associated paraneoplastic syndrome, largely due to the saturated presence of anti-cytokine autoantibodies (Burbelo et al., 2010; Ballman et al., 2022a). However, these patients were immunocompromised long before the clinical manifestation of immunodeficiency which left them vulnerable to present opportunistic infections, particularly recurrent and severe mucocutaneous candidiasis and pulmonary infections (Evoli and Lancaster, 2014). Hence, the development of targeted therapeutics that focus on the detection and prevention of early alterations in immune behaviour, such as T-cell hyperactivation, impaired regulation of positive selection and autoreactivity in the liver, are of profound clinical significance.

The current study’s revealed association between WHO classifications of tumour pathology and the recurrence of opportunistic infections post-thymectomy is not entirely surprising. WHO classification of thymic tumours has long served as an appropriate and reliable predictor of disease severity and overall survivalInfact, histological staging by WHO classification has served as a strong predictor of multiple criteria in clinical outcomes, post-thymectomy recovery and a moderate to strong predictor of thymic recurrence (Moran et al., 2012; Marx et al., 2015; Louis et al., 2021). Furthermore, the newly revised version of WHO on thymoma also accounts for immunohistological features of thymic pathology in its diagnostic criteria, such as further bifurcation of Type A and AB thymomas through perfuse presence of TdT+ T lymphocytes and distinctive cytokeratin networks (Suster and Moran, 2006; Marx et al., 2015). In addition, the clear links between the severity in the histology of thymic epithelial neoplasms by WHO classification and subsequent imbalance of immune regulation, particularly in the early phases of diagnosis, have been demonstrated in contemporary literature (Suster and Moran, 2006). Unsurprisingly, disruptions of key regulatory processes of the immune system lead to a substantially higher occurrence and recurrence of opportunistic infections, as evident in thymoma patients of the current study (Table 3). Although there are no studies to date that have heavily interrogated the association between WHO thymoma classification and recurrence of opportunistic infections, the results of the current study follow a plausible extension of the literature as it complements the existing links between high-grade thymic tumour histology leading to a highly dysregulated immune system, which further aids and abets the vulnerable immune system to opportunistic infections.

In addition, the current study’s derived association between increasing infiltration of thymic tumour based on the Masaoka staging and the increase in manifestations of paraneoplastic autoimmune syndromes follows the increasingly elucidated line of research on the impact of lymphatic dissemination and lymph node involvement (characteristic of high-grade Masaoka infiltration) and altered autoimmune response (Wilkins et al., 1991; Girard et al., 2009; Roden et al., 2015). Multiple studies have revealed the influence of metabolic derangements caused by lymphogenous metastasis of high-grade tumour infiltration to distal organs, characterized by Masaoka staging and the subsequent generation of chemokines and cross-reactions of tumour neoantigens, prompting the production of autoantibodies, and subsequent manifestations of paraneoplastic autoimmune syndromes such as MG, Good’s syndrome and Pure Red Cell Aplasia (Girard, 2019; Zhao and Rajan, 2019; Ballman et al., 2022). Furthermore, another contributing constituent of paraneoplastic autoimmunity is the presence of structural and functional similarities between autoantigens generated and expressed on distal organs and antigens expressed by thymic epithelial tumour cells (TETs), which further corroborates the link between the severity of tumour infiltration defined by Masaoka staging and increased presentations of paraneoplastic autoimmune syndromes (Radovich et al., 2018; Ballman et al., 2022).

The major findings of the current study, which were the two derived autoimmune associations discussed above, are further substantiated by the presence of a statistical association between multiple autoimmune comorbidities and the increased occurrence and recurrence of sub-severe and severe opportunistic infections. This was a particularly interesting finding as there was a nearly uniform presence of MG Anti-AChR autoantibody across the patient population in the current study, a predominance of severe infections across patients with multiple autoimmune comorbidities, and patients with the absence of Anti-AChR autoantibody did not develop any form of infection post-thymectomy. Although such an association merely recapitulates an interaction between two variables, that of infection and autoimmunity, it does, however, raise a consequential question about the nature of autoimmunity and vulnerability to opportunistic infections. Furthermore, given that paraneoplastic autoimmunity can either manifest as a mere symptom of thymic malignancy or can develop years and even a decade after diagnosis, post-thymectomy and or post-recurrence (Evoli and Lancaster, 2014; Lippner et al., 2019; Ballman et al., 2022) a greater interrogation by the contemporary literature on the temporal relationship between opportunistic infections and autoimmunity is needed. For instance, Myasthenia Gravis (MG) associated thymomas, such as the current study, are characterized by an overexpression of the neurofilament gene (NEF) which, in fact, share a substantial number of sequences with the gene coding for titin epitopes and acetylcholine receptors affiliated with MG (Radovich et al., 2018; Ballman et al., 2022). Hence, irrespective of the underlying shared mechanisms of autoimmunity and antibody expression profiles between thymic malignancies and paraneoplastic syndromes, novel therapeutics must further investigate the vulnerability and susceptibility to an infection created by such autoimmune conditions that are almost inevitably manifested in thymoma patients.

### Limitations and future directions

The current study shed light on multiple clinically relevant associations present in thymic malignancy patients suffering from a concomitant diagnosis of Myasthenia Gravis (MG). Furthermore, the current study established a much-needed frequency and type of multisystem autoimmune features and opportunistic infections prevalent in thymomas. This is particularly important as there is limited published literature on the epidemiology of thymomas, and much of the existing literature draws from small, retrospective, single-centre cohort studies and relatively few population-based studies (Engels, 2010; Kim and Thomas, 2015; Weis et al., 2015). However, the current study presented a few consequential limitations. First, there was no collected ethnic data on the thymoma patients of the current study, and many of the included patients were not diagnosed in Oxford. Rather, they underwent thymectomy at Oxford and received the rest of their treatment post-thymectomy at Oxford University Hospitals.

Although descriptive epidemiology of thymomas and thymic recurrence risk assessments are inconclusive and limited by the rare prevalence of such a condition, and most studies yield few clues on the aetiology of thymic malignancy, it has been well recorded that Asians and Pacific Islanders experience a higher incidence of thymoma, proposing the influence of genetic risk factors on the presentation of thymomas (Engels, 2010; Kelly et al., 2011). Furthermore, numerous studies on incidence trends of thymoma have recorded substantially lower incidence rates in Northern and Eastern Europe and highest in Central and Southern European countries (Siesling et al., 2012). The current study’s lack of such a demographic limit its external validity and the context and extent to which the derived immunological and clinical associations can be applied to aid preventive efforts and public health measures.

The highly heterogeneous treatments received by the patients prior to their thymectomy at Oxford pose an important limitation on the internal validity of the derived associations. This is largely because immunosuppressants, neoadjuvant radiotherapy and chemotherapy, and the time and thoroughness of the initial diagnosis, have a significant impact on the clinical and immunological presentations, conditions and disease trajectory associated with thymic malignancies. Another important limitation of the current study was the selection criteria of MG. Although all the patients in the current study presented with both a pathologically proven thymoma and MG, numerous patients developed MG and other paraneoplastic autoimmune syndromes long before their diagnosis of thymoma. This limits the extent to which conclusions can be drawn about the immunological cascades and antibody expression profiles post-thymectomy. Hence, in some patients, it may be unclear whether it was the initial MG diagnosis or the thymoma diagnosis that instantiated the immune imbalance, T-cell hyperactivation and immunological cascades responsible for the pathogenesis and clinical presentation of the disease.

Given the current study’s demonstrated association between histological criteria and comorbid paraneoplastic autoimmune presentations and recurrent opportunistic infections, future studies on thymic malignancies must further explore the link between autoimmunity, its associated antibody expression profiles and thymic tissue pathology. Furthermore, determining whether autoimmune presentations or opportunistic infections following tumour removal can herald the recurrence of thymic malignancies can be particularly insightful in shaping our understanding of how such a highly complex and heterogeneous condition reacts to immunological insults. In addition, given the overwhelming pathophysiological influence of T-lymphocyte hyperactivation and proliferation against thymus antigens and its subsequently induced immune imbalance, novel therapeutic interventions that generate an augmented T-lymphocyte reactivity toward TET cancer cells and enhance regulation of immune stimulatory checkpoints can serve as highly promising immunotherapeutics in the management of thymomas (Martin et al., 2015; Zhang and Chen, 2018; Ballman et al., 2022). Hence, a greater interrogation of novel immunotherapies such as Immune checkpoint inhibitors (ICIs), adoptive cell therapies (ACT) and cancer vaccines, which have substantially altered the treatment landscape for many other cancer diagnoses, could potentially alter our understanding of diagnosis, treatment and prevention strategies for such a highly complex and heterogeneous malignancy.

## Data Availability

All data produced in the present study are available upon reasonable request to the corresponding author.

